# Corneal cisterns as drainage aqueducts

**DOI:** 10.1101/2022.01.20.22268650

**Authors:** Shikha Gupta, Shivani Joshi, Karthikeyan Mahalingam, Abhishek Singh, Monika Arora, Viney Gupta

## Abstract

**Importance:** Anterior chamber paracentesis (ACP) is a known method of rapid reduction of IOP. The mechanism of how corneal fluid clears after ACP is not well described.

**Objective:** To determine the mechanism of instantaneous corneal clearing of edema after ACP.

**Design, setting, and participants:** This case series includes consecutive patients with high IOP and associated corneal edema, who presented to the Glaucoma Services of our tertiary care center. Pre- and post-intervention (ACP) ASOCTs were analyzed.

**Main outcome measures:** Resolution of corneal edema (clinical picture) and imaging of peripheral cornea and iridocorneal angle area.

**Results:** Ten consecutive patients who underwent an ACP were included in this study. The mean IOP pre-intervention was 48.4 ± 8.2 mmHg. ASOCT showed ill-defined conventional pathways of aqueous outflow, which were blocked by peripheral anterior synechiae in 4 eyes. Immediately post ACP, the IOP was 16.3 ± 5.6 mmHg (p < 0.001) and corneal edema resolved immediately. We found on ASOCT, that the intrastromal hypolucency of the peripheral cornea, (corneal cistern) which was limited in extent and U shaped, showed a posterior extension into the corneo-limbal junction to form well-defined anastomoses with the conventional aqueous drainage channels demonstrating a direct conduit for intracorneal fluid to egress. In 2/10 eyes, Schlemm’s canal opened intracamerally as well, suggesting direct reflux of intra-corneal fluid into the anterior chamber.

**Conclusion and Relevance:** After an ACP, in eyes with corneal edema to relieve IOP, we observed that the intracorneal cisterns undergo peripheral extension to drain corneal fluid through the conventional distal outflow channels.

## Introduction

High intraocular pressure (IOP) often leads to corneal edema by adversely affecting the active endothelial pump mechanism resulting in stromal edema, accentuated by cellular endothelial damage.^1–3^ It is well described that anterior chamber paracentesis (ACP) in patients with high IOP and corneal edema results in a rapid reduction in IOP as well as immediate corneal clearing.^4^ However, the mechanism is yet not fully elucidated. The current understanding is that endothelial pump becomes re-functional, thereby pumping the fluid from the cornea into the anterior chamber. We used high-definition Anterior segment optical coherence tomography (ASOCT) to observe the limbal aqueous drainage pathways to observe the fluid movement following an ACP.

## Methodology

Consecutive patients presenting to our tertiary care center with high IOP and associated corneal edema were enrolled. Ethical approval was given by the Ethics Committee of All India Institute of Medical Sciences, New Delhi. Written informed consent was obtained and the tenets of the Declaration of Helsinki were followed.

Patients underwent ophthalmic examination to ascertain visual acuity (VA in logMAR), IOP (Goldmann Applanation Tonometry), etc. We took anterior segment photos before and after the ACP (Eye Cap system, Haag Streit International, Switzerland). This was followed by imaging of the cornea and angle on high-resolution Spectral Domain ASOCT (Spectralis, Heidelberg Engineering, Germany). Immediately upon the corneal clearing, IOP was re-measured, and ASOCT was repeated within 15 minutes. We specifically examined the conventional aqueous drainage channels and their relationship to any corneal channels at the corneo-limbal junction.

## Results

A total of 10 patients were enrolled for this study with a median age of 46 years (Range: 12- 67). The mean IOP pre-intervention was 48.4±8.2 mmHg and post-intervention was 16.3±5.6 mmHg (p< 0.001) taken immediately post ACP. In all the patients, the corneal edema resolved immediately after ACP. Pre-intervention photograph of three eyes shows the presence of stromal edema with or without epithelial edema in all eyes (Fig 1a,b,c Pre). Following ACP, corneal edema resolved (Fig 1a,b,c Post).

**Figure 1.**
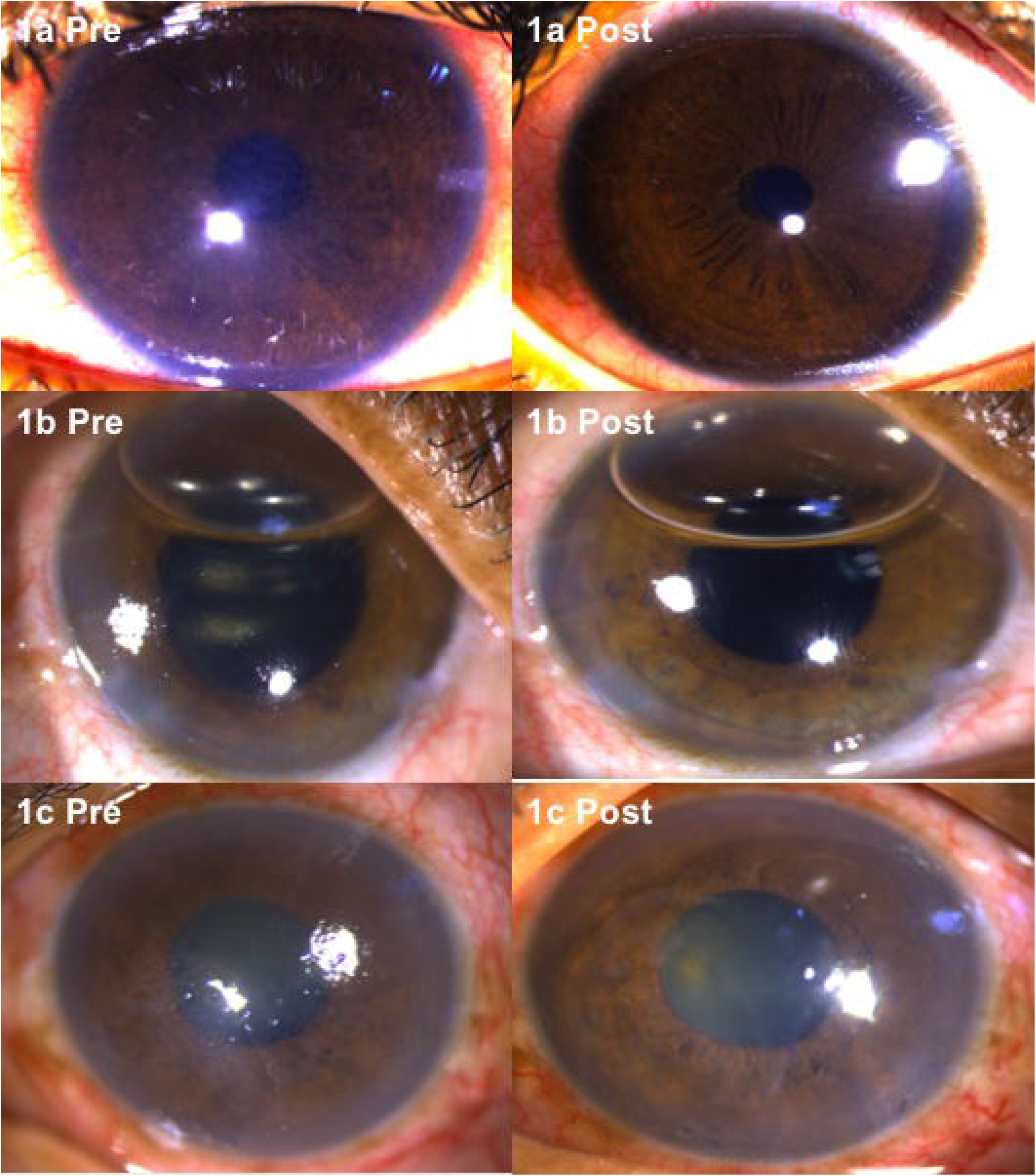
**a**,**b**,**c Pre:** Clinical picture of 3 representative patients at first presentation with corneal edema. **Figure 1a,b,c Post:** Anterior segment picture show immediate resolution of corneal edema after paracentesis in the same eyes. The details of the iris are now clear.

**ASOCT:** Pre-intervention ASOCT of the same 3 eyes as above mentioned shows a, well-defined, U-shaped, intracorneal cistern in the peripheral cornea (**Fig 2a Pre, arrow**). All these eyes has a non-discernable Schlemm’s canal (SC) (**Fig 2a,b**,**Pre**). In eyes with peripheral anterior synechiae (4/10; **Fig 2a, b, Pre**), none of the conventional aqueous outflow channels could be discerned including total absence of distal collector channels. Immediately following the intervention, the intracorneal cistern was found to dilate and extend posteriorly in all eyes. (**Figure 2a,b,c Post**) The distal collector channels also became well defined on IOP lowering and developed direct anastomoses with (**Figure 2**,**arrows**) the central corneal cistern suggesting a direct conduit for intracorneal fluid to egress through the conventional channels. In two eyes, the SC was seen to open intracamerally as well (**Figure2c Post, star**) suggesting that intra-corneal fluid can also reflux back into the anterior chamber.

**Figures 2.**
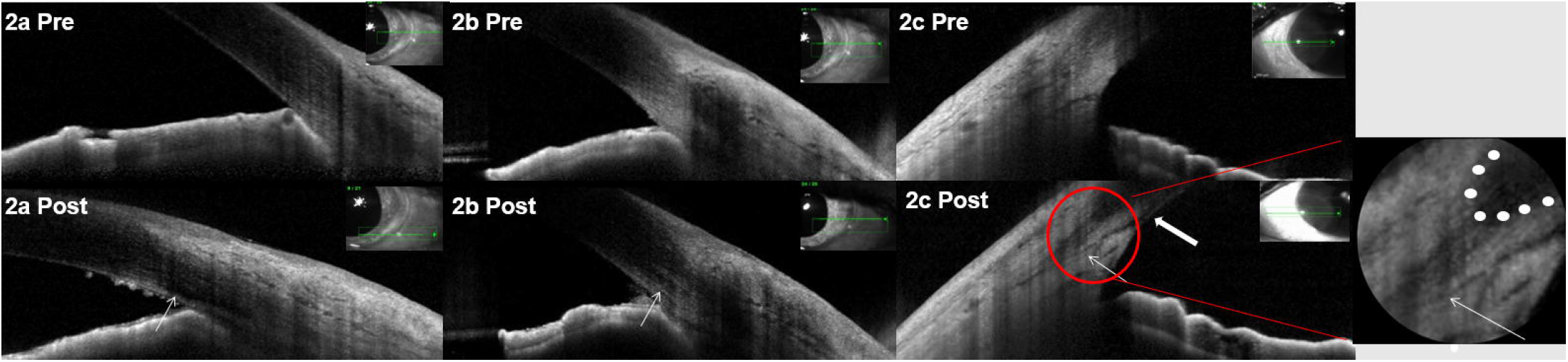
**a**,**b**,**c Pre**: Pre-intervention ASOCT images showing U shaped peripheral corneal cistern without any discernible Schlemm’s canal. **Figures 2 a,b,c Post**: Post-intervention images show the prominent collector channels (long arrow) having well-defined interdigitations with the corneal cistern that now extends further posteriorly. **Fig2a Post** shows collection of some inflammatory debris at the angle after ACP. **Fig 2c Post** shows drainage of Schlemm’s canal intracamerally (thick arrow). **Inset** shows magnified view of the posterior communication of the Schlemm’s canal with corneal cistern following intervention and dotted line shows the boundary of the corneal cistern in ‘U’ shape.

## Discussion

Anterior chamber paracentesis is used commonly to relieve raised IOP associated with corneal edema, especially in eyes with angle closure glaucoma. On imaging, we observed that high IOP was associated with collapsed outflow channels leading to impaired aqueous filtration through the conventional pathways. This has been described previously too.^4,5^ This may also explain absence of excessive fluid collection and of corneal haziness in eyes with not so high IOP elevation, when channels remain patent. However, with significant IOP lowering, the channels including the Schlemm’s canal (SC), were seen to open.

The anatomical characteristics of the peripheral cornea on ASOCT are not well defined. We used the term ‘corneal cistern’ to describe the peripheral “U” shaped intracorneal hypolucent area. This was also referred to as the lucid interval in a report.^5^ We believe that this cistern, acting as an aqueduct, facilitates the fluid movement from the edematous corneal stroma into the Schlemm’s canal and/ or the intrascleral collector channels. This fluid movement also explains how the corneal edema resolves in eyes that have peripheral anterior synechiae, by facilitating the movement directly from the cornea into the intra-scleral collector channels. Hence, the corneal cistern draining into the intra-scleral collector channels could play a pivotal role in immediate disappearance of corneal edema unlike the widely accepted mechanism of corneal endothelial pumps becoming re-functional.

Communication of the intra-corneal channels with conventional aqueous pathways has been suggested from the appearance of intra-corneal cavitation bubbles intracamerally during Femtosecond LASIK procedure, possibly by dissecting across the corneal lamellar planes peri-limbally and then entering intracamerally through the trabecular meshwork via Schlemm’s canal.^6–8^ Similarly, creation of lamellar intrastromal clefts in eyes with corneal hydrops in keratoconus aids in early resorption of corneal fluid as immediate entry of these pockets to outside is provided.^9^ Our study further strengthens this observation that limited corneal cisterns dilate and extend peripherally to drain any sequestered corneal fluid into the conventional aqueous outflow channels.

In a previous study, we have also shown that SC can open backward into the intracameral space both on in vivo imaging as well as on histopathology in fellow eyes of congenital glaucoma.^10^ Similar opening of SC intracamerally was seen in 2 eyes post ACP (**Fig 2c Post**), suggesting another route of clearing for corneal edema.

Our observations provide a mechanistic explanation to the instantaneous clearing of corneal edema with lowering of IOP, post ACP, which was till now, not well understood. Apart from this, it provides credence to alternative drainage of corneal fluid through the peripheral cistern and its communication with the conventional aqueous outflow channels.

## Data Availability

All data produced in the present study are available upon reasonable request to the authors

